# Effect of WASH/MNCH Integrated interventions on skilled birth attendance and incidence of neonatal Sepsis in Amuru District, Uganda; A quasi-experimental study

**DOI:** 10.1101/2022.09.29.22280511

**Authors:** Comfort Hajra Mukasa, Maureen Nankanja, Margaret Mugisa, Ojoro Valentine, Patrick Kagurusi

**Author notes:** **Correspondence** Comfort Hajra Mukasa,. **Contact information of co-authors** Comfort Hajra Mukasa. Maureen Nankanja Margaret Mugisa Ojoro Valentine Patrick Kagurusi.

## Abstract

**Background:** Evidence on the impact of Maternal Newborn and child health (MNCH) and Water, Sanitation and Hygiene (WASH) interventions on skilled birth attendance and neonatal sepsis remains unclear. We assessed the effect of WASH/MNCH integrated interventions on skilled birth attendance and incidence of neonatal Sepsis in a resource-constrained setting in Uganda.

**Methods:** A quasi-experimental study design was conducted in Amuru district. The package of interventions implemented included; training of health workers, facilitation of integrated outreach services, construction of WASH facilities, and health education of communities. A digitized structured questionnaire was used to obtain data on ANC and skilled birth attendance, WASH practices and prevalence of pneumonia and diarrhea among 466 expectant mothers and caretakers of under-fives at baseline, midterm and endline. Data on the incidence of sepsis, ANC and skilled birth attendance, and WASH status was also obtained from 6 healthcare facilities. A total of 12 key informant interviews and 12 Focus group discussions were also conducted. Data were imported into STATA 15 for analysis. Two sample tests of proportions were used to compare findings at baseline and endline. Qualitative was analyzed using thematic content analysis.

**Results:** There was a significant increase in the number of women delivering at the health facilities that were supported by the project from 41.4% at baseline to 63.0% at endline (p= <0.0001). There was a reduction in the incidence of neonatal sepsis from 0.6% to 0.2% (p = 0.0687), although the difference was not significant. There was an increase in the percentage of households with sanitation facilities and improved hygiene practices. Community-level findings also indicated a decline in cases of water-borne illnesses; cases of dysentery decreased from 10.0% at baseline to 0.6% at endline, cases of cholera decreased from 8.9% to 1.9% at endline, cases of typhoid decreased from 26.5% to 12.7% at endline.

**Conclusion:** This study revealed that integrated WASH/MNCH interventions can significantly increase ANC and skilled birth attendance, reduce incidences of neonatal sepsis, diarrhea, pneumonia, and other related diseases and improve WASH practices in communities. Significant improvements in WASH/IPC in the maternity wards and the capacity of healthcare workers to deliver clean and safe MNCH services can also be realized. We recommend the integration of WASH/MNCH interventions for projects aimed at improving skilled birth attendance and WASH practices and reduction of childhood infections.

## Background

Maternal and neonatal mortality remain serious public health concerns, especially in low-and middle-income countries (LMICs). In 2017, over 295,000 women worldwide died from preventable causes related to pregnancy and childbirth (WHO, 2022b). Over the past years, under-five mortality has substantially reduced, but neonatal mortality remains stagnated and high (Sharrow et al., 2022, UNICEF, 2021). Approximately 2.4 million neonatal deaths were reported in 2020 (Asiimwe et al., 2019, UNICEF, 2021). Sub-Saharan Africa (SSA) is disproportionately affected and carries the largest share of maternal (66%) and neonatal deaths (27 deaths per 1000 live births) compared to other regions (WHO, 2022a, WHO, 2022b, UNICEF, 2021). Similarly, in Uganda, recent estimates also point to high maternal and neonatal mortality rates of 375 deaths per 100,000 live births and 19 deaths per 1,000 live births, respectively (World Bank, 2022).

Nearly three-quarters of all maternal deaths are caused by haemorrhage, infections, pre-eclampsia and eclampsia, and complications from delivery and unsafe abortion (WHO, 2022b). In contrast, sepsis, pneumonia, tetanus, diarrhea, prematurity and birth asphyxia account for the majority of neonatal deaths (Kananura et al., 2016). Particularly, sepsis is responsible for 225,000 neonatal deaths globally per annum (Tumuhamye et al., 2020). Poor maternal and neonatal outcomes stem from a lack of quality care during pregnancy and childbirth, especially in LMICs (Dahab and Sakellariou, 2020) like Uganda. In addition, complications after childbirth such as infections have partly been attributed to poor hygiene and sanitation practices (WHO, 2019, Dahab and Sakellariou, 2020, Campbell et al., 2015).

Existing evidence shows that majority of the maternal and neonatal deaths can be averted with improved access to and utilization of skilled birth attendance at all levels (Stenberg et al., 2014, Wafula et al., 2021, Wilunda et al., 2015), and improved access to Water, Sanitation and Hygiene (WASH) at the healthcare facility and community levels (Benova et al., 2014a, Campbell et al., 2015). Adequate WASH services are a prerequisite for the delivery of quality MNCH services including hygienic birth practices such as hand washing by birth attendants, cleaning the maternal perineum, use of a clean birth surface, clean cutting and tying of the cord. WASH interventions can reduce neonatal sepsis deaths, neonatal tetanus deaths and maternal sepsis deaths (Blencowe et al., 2011, Pollard et al., 2013, WHO and UNICEF, 2021, Campbell et al., 2015). Despite this, many healthcare facilities (HCFs) and communities in LMICs lack basic WASH. Over 50% of the HCFs lack basic water sources, 26% lack hand hygiene services at points of care, 63% lack basic sanitation services, and 70% lack basic healthcare and waste management services (WHO and UNICEF, 2021). In addition, the Uganda Demographic Health survey estimated that only 74% of births are assisted by a skilled birth attendant, which still falls short of the 90% target (Munabi-Babigumira et al., 2019, UDHS, 2016).

Owing to the public health significance of maternal and neonatal mortality, countries adopted various strategies and committed to the UN Sustainable Development Goals (SDGs). SDG 3.1, particularly focuses global attention on the reduction of the maternal mortality ratio (MMR) to fewer than 70 maternal deaths per 100,000 live births by 2030 (Callister and Edwards, 2017). Furthermore, SDG 3.2 includes an ambitious target of “Ending preventable deaths of newborns and children under 5 years of age by 2030”, with all countries aiming to reduce neonatal mortality to at least as low as 12 per 1,000 live births and under-five mortality to at least 25 per 1,000 live births (WHO and UNICEF, 2020). Progress has been made over the years and maternal mortality has dropped substantially. However, the reduction is less than half the 6.4% annual rate needed to achieve the SDG global goal of 70 maternal deaths per 100,000 live births (UNICEF, 2021). In addition, the gains in the reduction in neonatal mortality have been slow over the years (UNICEF, 2021) and remain stagnated in countries like Uganda (UDHS, 2016).

Meeting national and global commitments needs a combination of strategies, including integration of Maternal, Newborn and Child Health (MNCH) and Water, Sanitation and Hygiene (WASH) interventions. Whereas there is consensus on the importance of WASH/MNCH for the prevention of maternal and neonatal mortality (Benova et al., 2014b, Delele et al., 2021), estimating the size of the effect has been more challenging (Benova et al., 2014b). Furthermore, there are few reports addressing the potential benefits and challenges of integrated programming, despite the conceivable linkages between WASH and MCH services. This study generated evidence on the effect of integrated MNCH /WASH interventions on MNCH outcomes to inform strategies that can be scaled up in similar settings for improved MNCH.

## Methodology

### Study design and setting

A quasi-experimental study design comparing non-randomly assigned intervention and control groups was conducted. Cross-sectional surveys employing both quantitative and qualitative data collection methods were conducted at baseline (pretest), midline and end line (posttest). The pre-intervention period (pretest) acted as the control. This study was conducted in Amuru District, Northern Uganda. Amuru district is located in Northern Uganda and is bordered by Adjumani district to the north, South Sudan and Lamwo district to the northeast, Gulu District to the east, Nwoya district to the south, Nebbi district to the southwest and Arua district to the west. The district was established by the Ugandan Parliament in 2006 as a breakaway from Gulu District and makes part of the Acholi sub-region. It has a total of 32 health facilities including 21 health center IIs, 10 health center IIIs and one Health center IV. The District is made up of 5 Lower Local Governments (4 Sub-counties and 1 Town Council). Other administrative units include 1 county, under the supervision of an Assistant Chief Administrative Officer. There are 32 parishes and 63 villages in Amuru District, with a total population of 186,696 people (Amuru District Local Government, 2021).

### Interventions

Interventions were delivered through the Total Health Project (THP) which was implemented by Amref Health Africa in Uganda in partnership with the Netherlands government. The project integrated WASH and MNCH interventions in four sub-counties of Amuru district including; Amuru, Pabbo, Attiak and Lamogi sub-county. From the four sub-counties, six parishes and six healthcare facilities were purposively selected to receive the interventions for a period of three years (2019-2021). These health facilities and communities were identified and selected under the guidance of the District Health Team for having the highest WASH and MNCH needs. The package of interventions implemented included the following:

1. On-job training and mentorship of health workers on current MNCH services
2. Facilitating integrated outreach services for health workers serving in the high-volume facilities
3. Installing water systems to provide running water in sluice rooms and maternity wards
4. Constructing lined pit latrines
5. Constructing post-delivery washrooms at health centers
6. Training community artisans to construct low-cost appropriate improved latrines.
7. Provision of essential services to six health facilities in the district
8. Training Village Health Teams (VHTs) to promote door-to-door health education using the new Community health workers (CHWs) guidelines.
9. Home improvement campaigns; triggering communities to improve their sanitation through Community Led Total Sanitation (CLTS) model and to facilitate follow-up missions of triggered communities.

These interventions were aimed at contributing to the overall outcomes namely; improved WASH practices, reduced incidence of neonatal sepsis, increased skilled birth attendance and reduced maternal and neonatal mortality in Amuru district. Specifically, the THP strengthened the capacity of skilled health workers to conduct clean and safe MNCH services, improved access to safe and clean MNCH services, improved sanitation and hygiene practices in households in selected sub-counties in Amuru district and promoted governance of WASH and MNCH services in the district.

### Study population and eligibility criteria

The study population included pregnant women, caretakers of children under five years, healthcare workers, health facility in charges and focal persons, community health workers and the district health team including health assistants and health inspectors. The district health team members were interviewed as key informants while pregnant women and caretakers of children under five were involved in the household surveys and focus group discussions (FGDs). Only expectant mothers or caretakers of children under-five aged 18 years and above were included in the study. Besides, we recruited only mothers/caretakers that had resided in the project areas for at least 6 months prior to the project. Households with an expectant mother or caretaker of a child/children under five who was critically ill or not in the right mental state to respond to questions were excluded.

### Data collection methods and tools

A household survey was conducted among expectant mothers, mothers/caretakers of children under five using a digitized structured questionnaire. The tool was used to obtain data on; the socio-demographic characteristics, history of ANC attendance, skilled birth attendance, WASH practices and prevalence of pneumonia and diarrhea at the community level. Health facility surveys were also conducted using digitized structured questionnaires to elicit data from healthcare workers on; the incidence of sepsis, diarrhea and pneumonia at the health facility, ANC attendance, skilled birth attendance, and WASH status of the facility. The RAs identified the respondents at the healthcare facilities, sought permission and consent and administered the questionnaires. An observational checklist was used to verify the state and functionality of visible aspects of WASH and MNCH in the households and health facilities. All surveys and qualitative interviews were conducted three times, at baseline, midline at 1 year after implementation, and the endline evaluation at 3 years. Qualitative interviews (Focus group discussions and Key informant interviews) were conducted using respective guides to explain the changes in key indicators. Each FGD consisted of homogeneous groups of between 8-12 women. Two RAs, a moderator and a note taker conducted the interviews.

### Sample size considerations and sampling technique

#### Quantitative component

##### Household survey sample size

The household sample size was calculated using the formula below;

n=N/ [1+N (e^2^)], where: e=0.046 (4.6%) is the desired 95.4% level of precision, N= Beneficiary households are estimated at 35,921 (Total number of Households in targeted sub-counties, n=required sample size. Therefore, a sample size of 466 pregnant women/children caretakers was targeted in each of the three surveys.

##### Health centre survey sample

Six high-volume healthcare facilities were selected for the interventions and all the 6 where were included in the three surveys. For each health facility, two respondents were purposively selected to respond to the MNCH and WASH questionnaire. These were selected based on their knowledge and experience of the intervention activities and implementation of WASH and MNCH activities in the health facilities. They included in-charges and program focal persons. The MNCH questionnaire was responded to by either the facility in charges, or MNCH focal person depending on the availability, while the WASH questionnaire was responded to by the facility in charges. The health inspector/assistant responded to the WASH questions.

##### Qualitative component

A total of 12 key informant interviews (KIIs) and 12 Focus group discussions (FGDs) were conducted. The sample sizes for both KIIS and FGDs were determined by thematic saturation. Key informants included; CAO/ District Planner/ Secretary for social services; Water User committee/ Health Unit Management committee member, Village Health Teams VHT/ Community Health Workers (CHWs), Project staff and in-charge of Health facilities. FGD participants included women/caretakers of children under five and expectant mothers. These were identified with help from the local council leader of the village.

##### Sampling technique

The THP was implemented in all the four sub-counties of Amuru district. The project targeted health facilities and specific parishes in each of the Sub-counties. The health facilities and parishes were purposively selected with the guidance of the district health team for having the highest WASH and MNCH challenges in the district. Proportionate sampling to size strategy was used to ensure representativeness of all sub-counties. The 466 expectant mothers/ children caretakers were selected proportionate to size for each sub-county, parish and village according to the respective household population. The village sample was further divided by 15 (minimum number of households per village) to yield sub-villages to be sampled in each village. In each sub-village, an average of 15 households with expectant mothers/mothers of children less than 5 years were selected using systematic sampling techniques. Where a selected household did not have an expectant mother/ child under five, it was skipped. The next household with the eligible participant was taken. In case the sampled household had more than one mother with a child under 5 years or more than one child under five years, only one was chosen using simple random sampling. The households were selected with help from the local council one (LC1) chairperson or his representative. The chairperson or their representative introduced the research assistant (RAs) to the household head from whom the RAs sought permission to talk with the expectant mother/caretakers of children under five years. Key informants that had worked in the project area for at least one year and interfaced with the intervention were purposively selected for the study.

##### Key indicators and measurement

The key outcome variables included ANC attendance, skilled birth attendance, incidence of sepsis, the prevalence of pneumonia, diarrhea and other related diseases and sanitation and hygiene practices of the mothers. Skilled birth attendance was defined as the proportion of women who delivered from the project healthcare facilities. This was recorded through a review of registers at the maternity wards in these facilities. The number of women attending ANC in each facility was also recorded and compared across the three surveys. Cases of sepsis were also recorded from health facility records and compared across the three surveys in the different sub-counties. Furthermore, the prevalence of pneumonia and diarrhea among children under five was measured by asking the mothers/caretakers if their household had experienced each of those infections in the past 2 weeks preceding the survey. We also assessed sanitation by establishing latrine coverage and status of open defecation in the target communities. Hygiene practices were measured by asking whether mothers/caretakers exhibited the following; washed hands with soap before feeding child or preparing food, washed hands with soap after using latrines, cleaned latrine every day, cleared bushes around the home, kept children’s cloths clean, keeping utensils clean, and disposed of children’s in the latrine and having a rack for utensils in the latrine

##### Data management and analysis

Data were uploaded onto a common server at the end of each field day. Data were checked for any errors and inconsistencies. At the end of each survey, data were exported to Microsoft Office Excel for cleaning then imported into STATA version 15 for analysis. Continuous variables were presented as means with standard deviations. Frequencies and percentages were used for the categorical variables. Categorical characteristics were compared between baseline, midline and end line data. Two sample test of proportions was used to compare findings at baseline and end line. A two-sided statistical test will be used with 95% confidence interval. For qualitative data, audio recordings were transcribed verbatim and then translated to English by a team of experts. Transcripts were read thoroughly to gain understanding of the context of each KI and FGD. Thematic content analysis was used to process all responses along with identification of relevant concepts and ideas found in the transcripts linked to the topics of inquiry. Relevant ideas were coded and categorized under specific themes. Emerging themes were included in the final matrix.

##### Quality control and assurance

Data was collected by research assistants who were supervised by the principal investigators. The data collectors were trained on data collection procedures and research ethics prior to start of the study, with refresher sessions organized when need arose. Data were checked at the end of each day by the principal investigators for any errors or inconsistencies. Only the study investigators, data manager, and authorized persons had access to the data files, and changes were only made with specific permission from the principal investigator.

##### Ethical statement

Ethical approval was obtained from the Mildmay Uganda Research Ethics Committee (MUREC) (MUREC-2021-72). The study was also registered with the Uganda National Council for Science and Technology. Administrative clearance was sought from the district authorities and the local leaders in the implementation parishes and villages. Informed consent was sought from the respondents prior to all interviews. We ensured that confidentiality was maintained.

## Results

### Socio-demographic characteristics of the respondents for the household survey

In total, 784 women (469 at baseline and 315 at endline evaluation) were included in the analysis. The average age of the respondents was 29 (±7.7) years at baseline and 29.6 (±7.7) at endline years. At baseline, about 35.8% of the respondents had no formal education compared to 15.0% at endline, 77.8% were catholic at baseline compared to 71.4% at endline, and 86.5% were married/cohabiting compared to 98.7% at endline (Table 1).

**Table 1:**
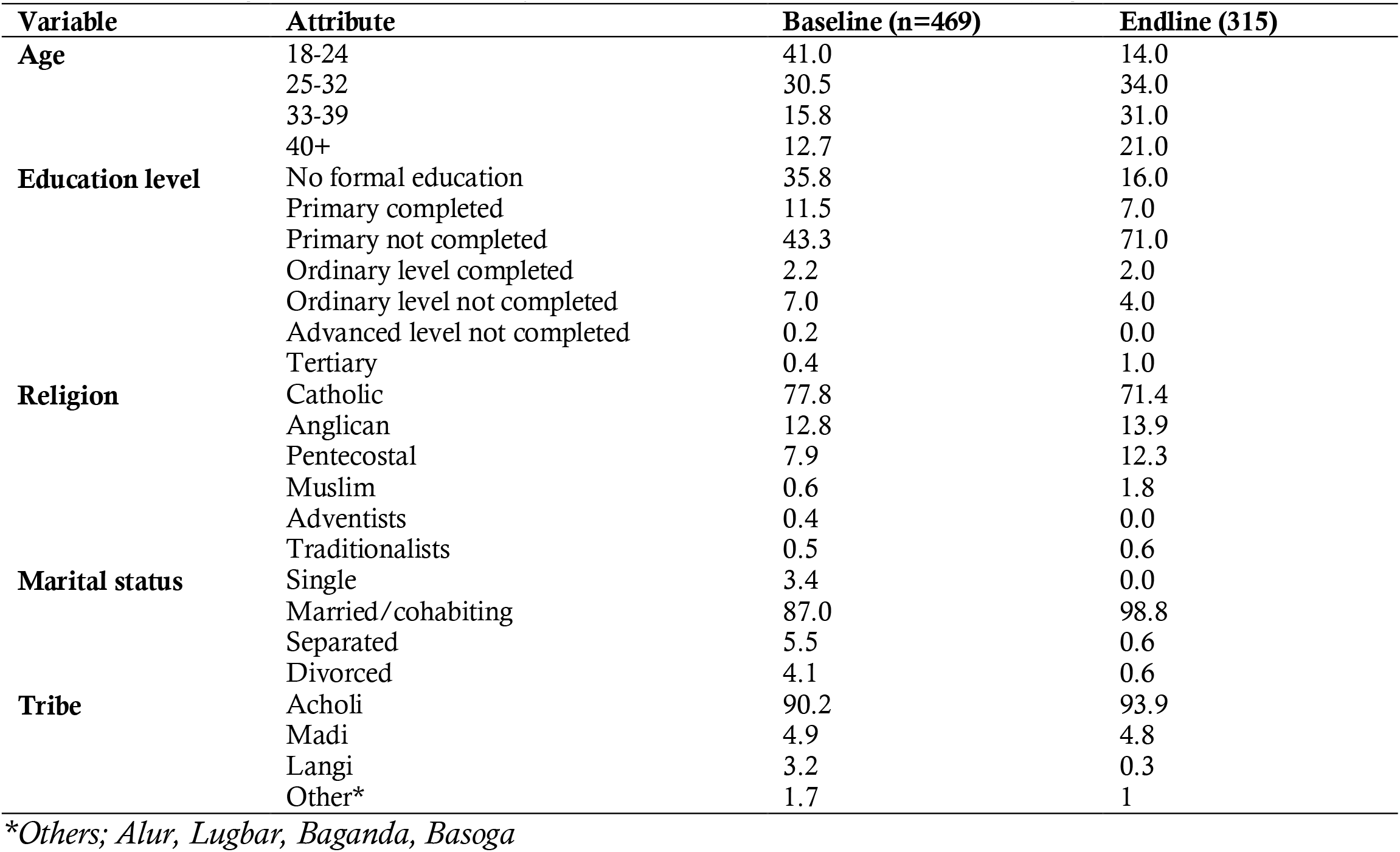
Socio-demographic characteristics of the household members in Amuru district, Uganda.

### Effect of the integration of WASH and MNCH interventions on ANC and skilled birth attendance in Amuru district

Findings from the endline evaluation showed a significant increase in the number of women delivering at the health facilities that were supported by THP in Amuru District. There was a significant increase in the percentage of deliveries at Olwal HC from 2.4% at baseline to 11% at endline (p = <0.001); from 4.2% to 12% at Kaladima HC (p = <0.001); from 3.0% to 6% at Atiak HC (p = <0.001); and 5.1% to 15% at Pabbo HC (p = <0.001) (Table 2). Endline evaluation data also showed a significant improvement in the percentage of pregnant women who attended their 4^th^ANC visit in the last one year preceding the survey (Figure 1).

**Table 2:** Skilled birth attendance compared between baseline, midterm review and endline evaluation in Amuru district.

| Health Center | Baseline evaluation |  |  | Midterm review |  |  | Endline evaluation |  |  | P-value |
| --- | --- | --- | --- | --- | --- | --- | --- | --- | --- | --- |
|  | Deliveries in HC | Pregnant women and women of reproductive age (14-49) | % Deliveries | Deliveries in HC | Total of pregnant women and women of reproductive age (14-49) | % Deliveries | Deliveries in HC | Total of pregnant women and women of reproductive age (14-49) | % Deliveries |  |
| Olwal | 120 | 5019 | 2.4 | 239 | 782 | 7.5 | 336 | 3169 | 11.0 | <0.001* |
| Bibia | 233 | 1983 | 11.7 | 166 | 586 | 7.1 | 209 | 2344 | 9.0 | <0.001* |
| Kaladima | 114 | 2683 | 4.2 | 74 | 610 | 3.0 | 286 | 2465 | 12.0 | <0.001* |
| Labongogali | 280 | 1868 | 15.0 | 257 | 580 | 9.6 | 263 | 2668 | 10.0 | <0.001* |
| Atiak | 219 | 7375 | 3.0 | 140 | 748 | 4.1 | 263 | 4201 | 6.0 | <0.001* |
| Pabbo | 291 | 5668 | 5.1 | 74 | 723 | 2.5 | 430 | 2942 | 15.0 | <0.001* |
| **Total** |  | 24596 | 41.4 |  |  |  |  | 17789 | 63.0 | <0.001* |

**Figure 1:**
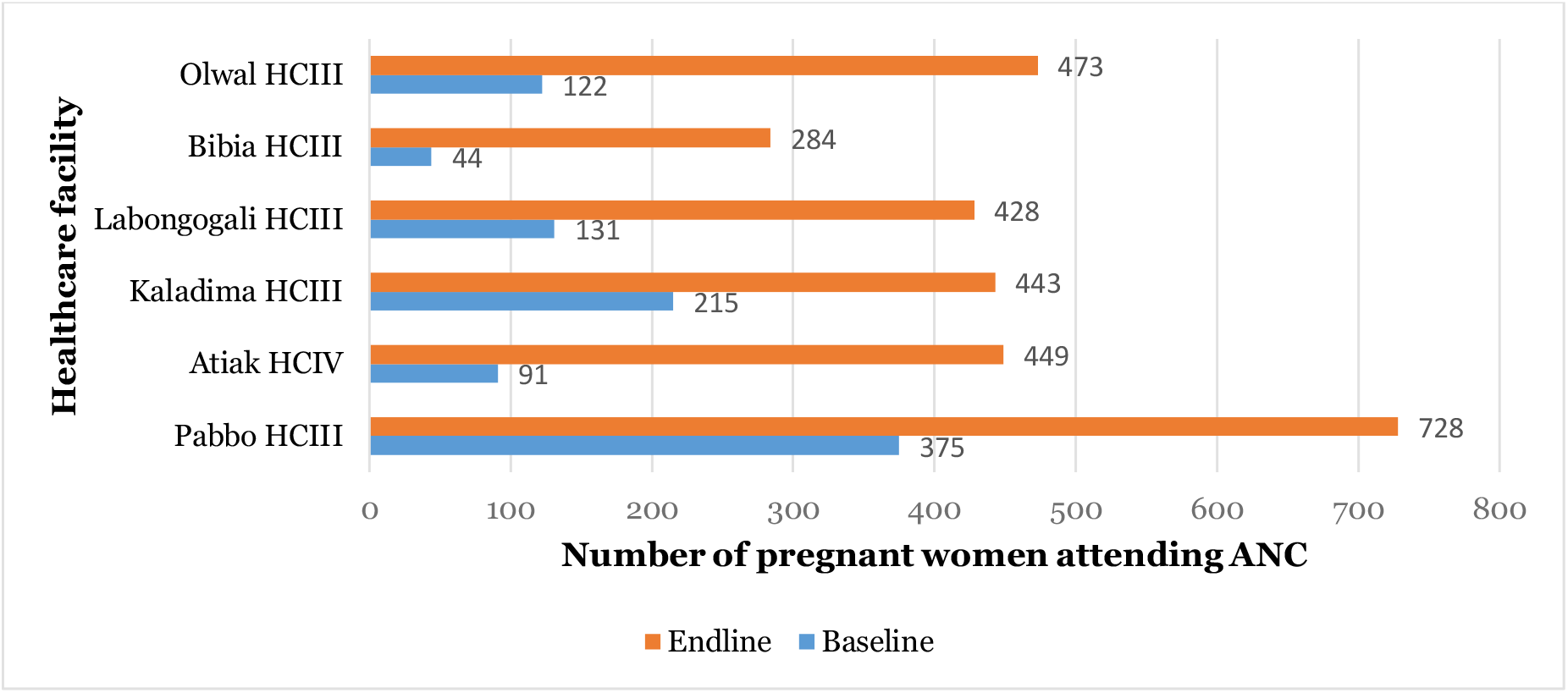
Comparison of the number of pregnant women attending the 4th ANC visit at baseline and endline in Amuru district.

### Effect of the integration of WASH and MNCH interventions on cases of neonatal sepsis, pneumonia and diarrheal diseases in Amuru district

Findings of EE revealed that integrating WASH and MNCH in the six health facilities reduced the incidences of neonatal sepsis from 0.6% at Baseline to 0.2% at EE (p = 0.0687). The difference was, however, not significant (Table 3). At healthcare facility level, there was also significant reduction in cases of Diarrhea, Pneumonia, and other related diseases in the targeted communities of Atiak, Lamogi, Amuru and Pabbo sub-counties as shown in the table 4 below. Community-level findings also indicated a decline in cases of water-borne illnesses; cases of dysentery decreased from 10.0% at baseline to 0.6% at endline (p=<0.001), cases of cholera decreased from 8.9% to 1.9% at endline (p=0.001), cases of typhoid decreased from 26.5% to 12.7% at endline (p=<0.001) (Figure 2). From the qualitative findings, expressed that the interventions under the THP such as: the renovation of the maternity units, provision of water, electricity and medical equipment, tools, drugs, etc., as contributed to a reduction in neonatal sepsis.

**Table 3:** Shows the number of neonates presented and cases of sepsis recorded at project health facilities that were supported by the THP:

| Health Facility | Baseline |  |  |  | Mid-Term Review |  |  |  | End-line |  |  |  |
| --- | --- | --- | --- | --- | --- | --- | --- | --- | --- | --- | --- | --- |
|  | Neonates | Male with Sepsis | Female with Sepsis | Total | Neonates | Male with Sepsis | Female with Sepsis | Total | Neonates | Male with sepsis | Female with sepsis | Total |
| Olwal HC III | 120 | 0 | 0 | 0 | 197 | 0 | 0 | 0 | 336 | 0 | 0 | 0 |
| Bibia HC III | 233 | 2 | 3 | 5 | 122 | 0 | 0 | 0 | 209 | 1 | 0 | 1 |
| Labongogali HC II | 279 | 0 | 0 | 0 | 334 | 0 | 0 | 0 | 321 | 0 | 0 | 0 |
| Kaladima HC III | 114 | 1 | 0 | 1 | 123 | 2 | 1 | 3 | 286 | 0 | 0 | 0 |
| Atiak HC IV | 219 | 0 | 0 | 0 | 125 | 1 | 0 | 1 | 264 | 2 | 0 | 2 |
| Pabbo HC III | 291 | 1 | 0 | 1 | 229 | 0 | 0 | 0 | 430 | 0 | 0 | 0 |
| All HCs | 1256 | 4 | 3 | 7 | 1,097 | 3 | 1 | 4 | 1,845 | 3 | 0 | 3 |
|  |  |  |  | 0.60% |  |  |  | 0.40% |  |  |  | 0.20% |

**Table 4:** Cases of diarrhea, pneumonia, and other related diseases in the targeted communities of Amuru district in the previous 6 months preceding the surveys.

| Sub-county | Baseline |  |  | End-line |  |  |
| --- | --- | --- | --- | --- | --- | --- |
|  | Diarrhea | Pneumonia | Others | Diarrhea | Pneumonia | Others |
| Amuru | 25 | 58 | 6 | 3 | 0 | 3 |
| Atiak | 18 | 64 | 3 | 6 | 3 | 2 |
| Lamogi | 20 | 62 | 8 | 2 | 2 | 5 |
| Pabbo | 23 | 66 | 6 | 2 | 1 | 1 |

**Figure 2:**
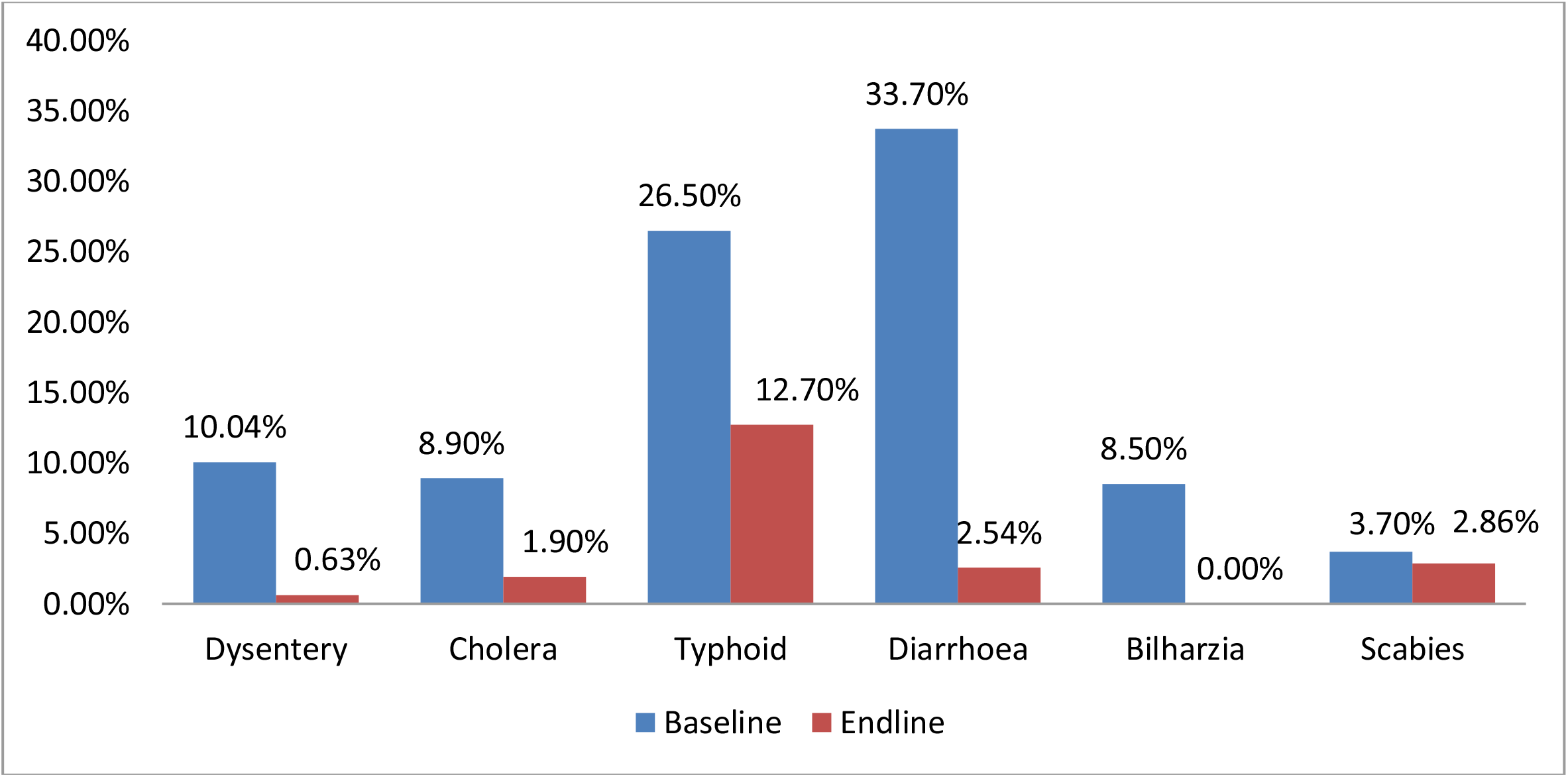
Prevalence of waterborne illnesses in targeted households in Amuru district 2 weeks preceding the survey.

> *The infrastructural development was the main reason for reducing neonatal sepsis. They connected water to the ward. The transmission mechanism behind neonatal sepsis is poor hygiene in the ward. This poor hygiene needs a lot of water for cleaning. It needs constant cleaning. When there is no water that is when you have bacteria which cause those infections like neonatal sepsis. Those areas have been addressed by THP. I have reported any new cases on neonatal sepsis. They have not reported to me. And also when I look at my statistics, neonatal cases are not seen… (****Key informant Interview 1***).

> *The situation of maternity units was very bad in terms of hygiene and sanitation and this was leading to high rate of neonatal sepsis and other infection among children and mothers before the implementation of THP by Amref Health Africa. There was no water in the maternity units and provision of running water and other supplies in contributed greatly towards improvement of hygiene and sanitation in the maternity units and reduction in the rate of neonatal sepsis situation was very bad and this was causing diseases… (****Focus Group Discussion Sub County 1***).

### Effect of the integration of WASH and MNCH interventions on sanitation and hygiene practices among households in Amuru district

Endline evaluation indicated that THP has greatly improved sanitation and hygiene practices at the household levels in Amuru District with special focus to children under five years of age and expectant mothers. This was attested by significant reduction in open defecation in the communities that were supported by THP and adoption of good hygiene practices in the communities as elaborated below.

### Change in latrine coverage and open defecation in the targeted communities in Amuru district

The highest percentage point increase in households where there was no observed open defecation was noted in Amuru (50.3%), followed by Pabbo 49.9%), Atiak (44.8%), and Lamogi (36.1%). All increments were statistically significant at 5%, implying that the Community-Led Sanitation activities significantly reduced open defecation and improved the sanitation situation in households in the project intervention areas (Table 5). The study also registered an increase in the percentage of households with sanitation facilities (Figure 3).

Qualitative findings also affirmed that there was a reduction in the practice of open defecation in the project area. Respondents reported that Seventeen 63% (17/27) of the villages were declared free of open defecation at the endline evaluation. Furthermore, the project called for continuous sensitization and monitoring of hygiene and sanitation practices, which also contributed to the improvement.

**Table 5:** Prevalence of open defecation in the project area at baseline and endline.

|  | Baseline (n=469) | Endline (n=315) | Difference |
| --- | --- | --- | --- |
| **Amuru** | 45.2% | 92.8% | +47.6% |
| **Atiak** | 49.3% | 94.1% | +44.8% |
| **Lamogi** | 59.3% | 95.4% | +36.1% |
| **Pabbo** | 55.1% | 100.0% | +44.9% |

**Figure 3:**
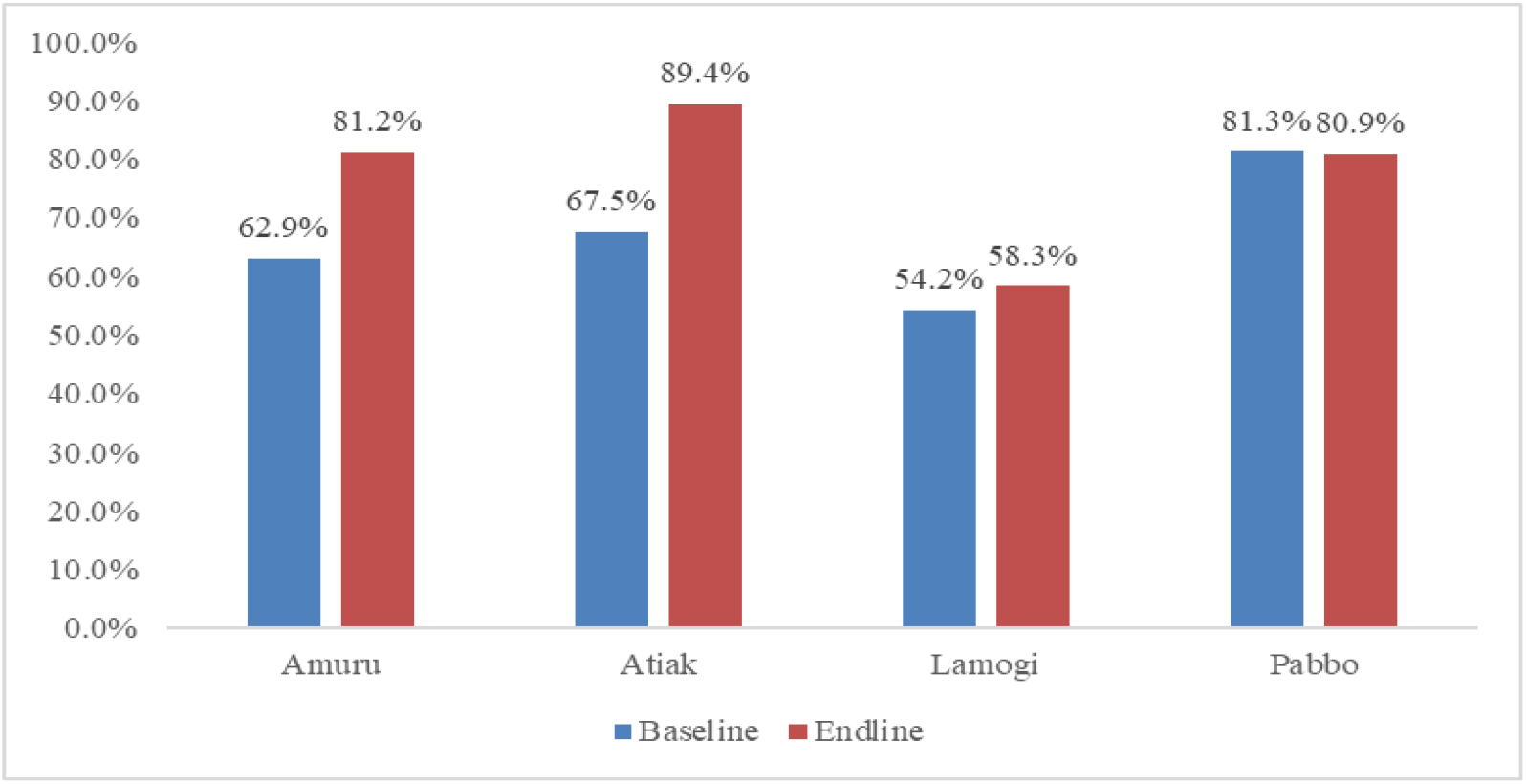
A bar graph showing Percentage of Households with Latrines in the Sub-Counties of Amuru, Atiak, Lamogi and Pabbo at baseline and endline.

> *There has been significant improvement in hygiene and sanitation among households with mothers and children under 5 and the general community members because open defecation has reduced and all HHs in the targeted villages of THP interventions have been using hand washing facilities and other hygiene and sanitation practices. We worked in 27 targeted communities. Seventeen (17) were declared open defecation free. Ten (10) were near. This means open defecation is still there but reduced in the villages where THP interventions have been implemented… (****KII 5***).

> *In the targeted households, most families that used to ease themselves in the bush have Latrines in their homes now. Currently, we have registered up to 17 villages that are Open Defecation Free. This is a tremendous change because it has not been easy for local government to achieve ODF. And with support from Amref Health Africa some villages were declared free and they have maintained that condition. They have not gone back to open defecation (OD) as it used to be. However, in some areas where AMREF has not visited, the practice still exists. Of course the target was 27 villages. We achieved 17 which were declared ODF. There was some more which should have emerged but since the project ended, we shall try to work around. There were promising villages to be declared free. We shall continue as local government to have them declared… (****KII 3***).

> *AMREF has been sensitizing communities…this has contributed to a great improvement in hygiene in our families. Community members have latrines. There is no open defecation. AMREF worked with the local leaders to enforce hygiene in the community as a strategy for promoting hygiene. All men with more than one wife were required to construct a latrine for each wife. Ten community members from the village were selected to carry out monitoring of hygiene situation. This contributed greatly towards improvement in hygiene… (****FGD 2***).

### Hygiene practices in the project communities in Amuru district

There were significant improvements in the hygiene and sanitation practices in sub-counties. Overall, there was significant improvement in the eleven areas that were assessed: Washing hands with soap before feeding a child or preparing food improved from 41% at baseline to 59% at EE (p=<0.001), washing hands with soap after using latrine improved from 31.8% at baseline to 59.0% at EE (p=<0.001), Cleaning the latrines every day improved from 29.8% at baseline to 60% at EE (p=<0.001), Keeping water and soap for washing hands at latrine improved from 16.6% at baseline to 43% at EE (p=<0.001), and Clearing bushes around the home improved from 49.2% at baseline to 60% at EE (p=<0.001) among others (Table 6).

**Table 6:** Showing hygiene practices at baseline and endline evaluations of the THP in Amuru district, Uganda.

| Indicator | Baseline (n= 469) % |  |  |  |  | Endline (n= 315) % |  |  |  |  | P-value |
| --- | --- | --- | --- | --- | --- | --- | --- | --- | --- | --- | --- |
|  | Amuru | Atiak | Lamogi | Pabbo | Total | Amuru | Atiak | Lamogi | Pabbo | Total |  |
| % of HHs washing hands with soap before feeding a child or preparing food | 40.0 | 30.8 | 43.1 | 50.9 | 41.0 | 53.0 | 47.0 | 67.0 | 69.0 | 59.0 | <0.001* |
| % of HHs washing hands with soap after using Latrine | 24.2 | 24.8 | 31.0 | 49.1 | 31.8 | 57.0 | 53.0 | 56.0 | 69.0 | 59.0 | <0.001* |
| % of HHs cleaning the latrines every day | 26.7 | 30.8 | 29.3 | 33.0 | 29.8 | 47.0 | 63.0 | 61.0 | 69.0 | 60.0 | <0.001* |
| % of HHs keeping water and soap for washing hands at latrine | 16.7 | 14.5 | 15.5 | 19.8 | 16.6 | 53.0 | 33.0 | 28.0 | 56.0 | 43.0 | <0.001* |
| % clearing bushes around the home | 49.2 | 53.0 | 52.6 | 41.5 | 49.2 | 67.0 | 63.0 | 44.0 | 67.0 | 60.0 | 0.0030 |
| % avoiding stagnant water around the home | 16.7 | 11.1 | 35.3 | 13.2 | 19.2 | 30.0 | 20.0 | 17.0 | 31.0 | 24.0 | 0.1065 |
| % of HHs keeping the children's cloths clean | 39.2 | 47.9 | 43.1 | 38.7 | 42.3 | 60.0 | 50.0 | 33.0 | 31.0 | 43.0 | 0.8459 |
| % keeping utensils clean | 43.30 | 45.3 | 39.7 | 46.2 | 43.6 | 80.0 | 63.0 | 44.0 | 69.0 | 64.0 | <0.001* |
| % disposing off children's faeces in the latrine | 18.2 | 19.3 | 21.7 | 20.3 | 20.3 | 57.0 | 20.0 | 39.0 | 28.0 | 36.0 | <0.001* |
| % of HHs having a rack for utensils in the home | 5.8 | 7.7 | 9.5 | 8.5 | 7.8 | 23.0 | 17.0 | 17.0 | 28.0 | 21.0 | <0.001* |

*\*p-value less than 5%*

### Improvement in WASH/IPC in the maternity wards at the target healthcare facilities

There was an improvement in the WASH status of the project healthcare facilities due to infrastructural developments. Respondents reported an improvement in status of WASH/IPC as a result of construction of placenta pits, provision of safe water, and provision of medical equipment like sterilization equipment, delivery pacts, and improved lighting.

> *…There has been a change in hygiene due to the construction of a placenta pit. Also, the presence of water addressed the challenge of looking for water off facility premises. Sterilization of instruments has led to reduction in the rate of neonatal sepsis since there is a good sterilizer and the instruments were boiled in the past. This is a great change brought by AMREF*… (***KII 11***).

> ..*Since there was no power in the maternity ward, we used to conduct delivery at night using torches but now we have solar lights and this has been a great change. Pregnant mothers used to pump water from the borehole but now the solar powered water pump enables water to flow direct from the tank to the maternity. This is a great change since it has improved the hygiene and reduction in the neonatal sepsis… (****KII 7***).

> *The condition of maternity ward was very bad before AMREF renovated. There was no water and the hygienic conditions were poor. AMREF provided Flip charts for educating pregnant women at the health facility. Medical personnel were trained in different areas including how to handle mothers. Quality of services provided by medical personnel has improved…* ***(KII 12)***

### Capacity of health workers to deliver clean and safe MNCH services in Amuru District

Qualitative findings also indicated an improvement in the capacity of healthcare workers in relation to the delivery of clean and safe MNCH services in Amuru district. Over 25 healthcare workers received training on customer care, provision of basic obstetrics, antenatal care, management of maternal complications and neonatal complications during delivery like birth asphyxia and obstructed labor, which greatly improved the quality of care.

> *We were trained on customer care, managing complications during delivery, handling pregnant mothers, management of obstructed labour, provision of basic obstetrics and emergency care of mothers in labour, resuscitation of babies, provision of antenatal and postnatal care as well as on job coaching and mentoring using senior health practitioners from outside Amuru District. This significantly contributed to enhanced capacity of skilled health workers to deliver clean and safe maternal new-born and child health services and improvement in the quality of service delivery… (****KII 3)***.

> *…Training which actually took place was expected to improve the mind-set, behaviours and attitudes of midwives on how to treat mothers. It’s all known throughout this country that midwives are rude. Midwives beat mothers, midwives abuse mothers so under this project they allocated some funding…We brought some senior midwives from Kampala to talk to these midwives… so that they can improve on their attitudes towards mothers…I think it was a very important training that needs to be emphasized*…*there was also equipment that were given under this project, diagnostic equipment, liquid soap for service delivery. They also supported my office with support supervision, going out in the field to see what is taking place…* ***(KII 1)***.

## Discussion

This study assessed the effect of WASH/MNCH integrated interventions on skilled birth attendance, the incidence of childhood infections and WASH practices in Amuru District, Uganda. The findings indicated that integrated WASH/MNCH interventions can significantly increase ANC and skilled birth attendance, reduce incidences of neonatal sepsis, diarrhea, pneumonia, and other related diseases and improve WASH practices in communities. Analyses also indicated significant improvements in WASH/IPC in the maternity wards at the supported healthcare facilities and the capacity of healthcare workers to deliver clean and safe MNCH services in Amuru District.

Overall, the number of women delivering at healthcare facilities supported by the project significantly increased between baseline and endline evaluations. This achievement can be attributed to the improvement in the quality of healthcare offered at these facilities and WASH status. Improvement in the quality of healthcare can be explained by the training held with the healthcare workers in these facilities. The training focused on enabling health workers to offer clean and safe MNCH services such as basic obstetrics services, antenatal care, and management of maternal complications and complications during delivery. The training also emphasized the mindset and behavior change of the health workers to make them more friendly and cautious towards pregnant women and mothers seeking MNCH services. Indeed, mothers who accessed maternal services at the THP-supported healthcare facilities expressed satisfaction with the quality of antenatal care, post-natal care and general maternity services received. Quality of basic maternal care has previously been associated with skilled birth attendance (Kruk et al., 2016, Peet and Okeke, 2019). In addition, the THP improved access to WASH services and renovated maternal wards in these facilities. Provision of amenities such as water, sanitation, electricity and other supplies has been linked with skilled birth attendant’s ability to offer quality care (Munabi-Babigumira et al., 2017, Mbonye and Asimwe, 2010). The study also revealed an increase in the percentage of pregnant women who attended their 4th ANC visit. It is likely that these women continuously noticed changes in the quality of services and infrastructure which informed their choice at the time of delivery. It is also probable that these women recommended skilled birth attendance to their peers. According to Nahar et al. (2022) women who utilized more than four ANC visits showed a greater tendency to use skilled birth attendants during childbirth than their counterparts. The findings altogether indicate a need to continuously improve the quality of care at health facilities in the dimensions of effectiveness, safety and responsiveness or patient-centeredness.

We also noted that integrating WASH and MNCH interventions decreased the incidence of neonatal sepsis at healthcare facilities and significantly reduced the incidences of diarrhea, pneumonia, and other related diseases in the targeted communities. Neonatal sepsis is a common condition in healthcare facilities, especially in LMICs and has been associated with poor WASH conditions. Improving WASH and IPC services through the renovation of the maternity units, construction of hygiene infrastructure, and provision of water, electricity and medical equipment at the THP-supported facilities therefore must have led to the reduction in the incidence of these infections recorded at the healthcare facilities. This finding is supported by existing literature. In order to prevent infections and provide quality care, healthcare facilities need to have a safe and accessible water supply; clean and safe sanitation facilities, hand hygiene facilities at points of care and sanitary facilities; and appropriate waste disposal systems (CDC, 2020). It has also previously been documented that energy (electricity) is an enabler of better health service delivery and universal health coverage (WHO, 2014). It is also worth noting that there were no other interventions ongoing in these facilities during this period to confound the outcomes. Therefore, it is warranted for authorities to improve access to water, sanitation services and energy in healthcare facilities to reduce the incidence of infectious diseases.

The endline evaluation further indicated great improvement in the sanitation and hygiene practices at the household level, attested by the significant reduction in open defecation and adoption of good hygiene practices in the communities supported by the THP. Significant improvements were recorded in practices such as washing hands with soap at critical moments including before feeding a child, preparing food and after using a latrine, as well as daily cleaning the latrines, availing water and soap for washing hands at the latrine and clearing bushes around the home. These changes can be attributed to the efforts of the VHTs aimed at improving WASH conditions in these communities. The THP, through the VHTs, implemented sensitizations of community members on sanitation and hygiene to enlighten them on the nexus between their practices, the environment and the incidence of water-related diseases. Sensitizations and mindset change have been reported to positively influence WASH practices in communities (URC, 2021, USAID, 2017, IRC, 2016). The VHTs also implemented effective outreach and advocacy programs to increase the construction of pit latrines and uptake of good hygiene and sanitation practices. This study indeed found an increase in latrine coverage in the supported communities. The project further rehabilitated water sources in these communities leading to easy access to safe water. It is probable and supported by literature (USAID, 2017) that the reduction in incidences of water-borne diseases, sepsis and other hygiene-related infections discussed above was in part contingent on these community interventions. It is therefore important to promote better WASH practices in communities through a combination of strategies including communication for behaviour change and the necessary WASH infrastructural improvements to eliminate barriers.

### Strengths and limitations

To the best of our knowledge, this may be the first study that assessed the impact of the integration of MNCH and WASH interventions on MNCH outcomes in Uganda. Our study was complemented by qualitative data collection methods which provided explanations for some of the observed changes. However, the study is not free of limitations. Some of the outcomes of the study (Sanitation and hygiene practices) were assessed through self-report, and thus the study was subject to social desirability bias. We, however, minimized this by assuring participants of privacy and confidentiality of their responses. We also used a quasi-experimental study design which may not have taken into account confounders that are often best handled through randomization. Because of the study design used, it was difficult to confidently attribute the changes seen in the community only to the project interventions. However, to the best of our knowledge, no other partners were present and offering similar components of the interventions within the project period. We recommend that a more rigorous study design such as an experimental study design is used to ascertain the impact.

## Conclusion

Integrated WASH/MNCH interventions can significantly increase ANC and skilled birth attendance, reduce the incidence of neonatal sepsis, diarrhea, pneumonia, and other related diseases and improve WASH practices in communities. Significant improvements in WASH/IPC in the maternity wards and the capacity of healthcare workers to deliver clean and safe MNCH services can also be realized. We, therefore, recommend the integration of WASH/MNCH interventions for projects aimed at improving quality of care, skilled birth attendance, reduction of childhood infections and improvement in WASH practices.

## Data Availability

All data produced in the present study are available upon reasonable request to the authors

## Acknowledgements

We would like to thank the study community for sparing their time to participate in the study. Credit also goes out to the research assistants for their tremendous effort and dedication during data collection.

## Funding

Funding for this study was provided by Amref flying Doctors in Netherlands. The funding body did not play any role in the design of the study and collection, analysis, and interpretation of data and in writing the manuscript.

## Author’s contribution

PK, CHM, MM and MN conceptualized the study, participated in data collection, analysis and drafting the manuscript. OV participated in the analysis and drafting of the manuscript. All authors read and approved this manuscript before submission to this journal.

## Consent for publication

Not applicable.

## Availability of data and materials

The data used for this manuscript is available from the corresponding author on reasonable request.

## Competing interests

The authors declare that they have no competing interests.

## Notes

### Competing Interest Statement

The authors have declared no competing interest.

### Funding Statement

This study was funded by Amref Flying Doctors Netherlands

### Author Declarations

Mildmay Uganda Research and Ethic Committee (MUREC)

